# A retrospective longitudinal study of adenovirus group F, norovirus GI and GII, rotavirus, and enterovirus nucleic-acids in wastewater solids at two wastewater treatment plants: Solid-liquid partitioning and relation to clinical testing data

**DOI:** 10.1101/2023.08.29.23294748

**Authors:** Alexandria B. Boehm, Bridgette Hughes, Dorothea Duong, Niaz Banaei, Bradley J. White, Marlene K. Wolfe

## Abstract

**Background:** Enteric infections are important causes of morbidity and mortality, yet clinical surveillance is limited. Wastewater-based epidemiology (WBE) has been used to study community circulation of individual enteric viruses and panels of respiratory diseases, but there is limited work studying concurrent circulation of a suite of important enteric viruses.

**Methods:** A retrospective WBE study was carried out at two wastewater treatment plants located in California, United States. Using droplet digital polymerase chain reaction (PCR), we measured concentrations of human adenovirus group F, enteroviruses, norovirus genogroups I and II, and rotavirus nucleic-acids in wastewater solids two times per week for 26 months (n=459 samples) between 2/1/21 and 4/14/23. A novel probe-based PCR assay was developed and validated for adenovirus. We compared viral nucleic-acid concentrations to positivity rates for viral infections from clinical specimens submitted to a local clinical laboratory to assess concordance between the data sets.

**Findings:** We detected all viral targets in wastewater solids. At both wastewater treatment plants, human adenovirus group F and norovirus GII nucleic-acids were detected at the highest concentrations (median concentrations greater than 10^5^ cp/g), while rotavirus RNA was detected at the lowest concentrations (median on the order of 10^3^ cp/g). Rotavirus, adenovirus group F, and norovirus nucleic-acid concentrations were positivity associated with clinical specimen positivity rates. Concentrations of tested viral nucleic-acids exhibited complex associations with SARS-CoV-2 and other respiratory viral nucleic-acids in wastewater, suggesting divergent transmission patterns.

**Interpretation:** This study provides evidence for the use of wastewater solids for the sensitive detection of enteric virus targets in WBE programs aimed to better understand the spread of enteric disease at a localized, community level without limitations associated with testing many individuals. Wastewater data can inform clinical, public health, and individual decision making aimed to reduce transmission of enteric disease.

## Introduction

Infectious diseases are leading causes of morbidity and mortality^1^, and climate change may exacerbate the spread of critical and emerging diseases^2^. Diarrheal illness, in particular, is a leading cause of morbidity and mortality especially for children^3^. Enteric viruses, including human noroviruses, adenoviruses, enteroviruses, and rotavirus, are responsible for a large percentage of diarrheal illnesses^4,5^. Unfortunately, there is limited data on the incidence and prevalence of enteric viral infections and their etiologies in most of the world. Data that do exist are biased to severe cases, or individuals with comorbidities, and are often limited to syndromic data on GI symptoms that are non-specific. This limits the ability to inform prevention and response activities, including vaccine development, vaccination campaigns, pharmaceutical and non-pharmaceutical interventions.

Wastewater-based epidemiology (WBE) is emerging as a tool for assessing population-level disease occurrence. Wastewater contains excretions from individuals that enter drains and toilets including feces, urine, sputum, mucus and saliva. When individuals are infected by a pathogen, that pathogen or its components (nucleic-acids and proteins) can be excreted and enter wastewater. When wastewater is collected at a wastewater treatment plant (WWTP), it represents a composite sample of all the people in the sewershed (anywhere from ∼10^4^ - 10^6^ individuals). This includes even infected and shedding individuals who are asymptomatic or mildly symptomatic and those who do not seek or receive medical care. Results regarding presence and concentration of infectious disease targets can be available 24 hours after sample collection. Therefore, WBE data can be complementary to clinical data by providing rapid results at a community scale and may even provide early warnings of disease spread. Work to date suggests that wastewater monitoring are related to community disease levels and can be used to detect occurrence of important respiratory^6,7^, enteric^8,9^, and emerging and outbreak infections^10,11^.

Wastewater is a mixture of liquids (water, urine, and other liquids), and solids or particles (feces, food, cells, and other material). Viruses tend to partition to the solids in wastewater, where their concentrations can be orders of magnitude higher than in the liquid on a mass equivalent basis^12,13^. It has been proposed that using wastewater solids can provide sensitive measurements for WBE applications, but the affinity of enteric viruses to wastewater solids in WBE applications has not been previously described.

The goal of this study is to test the utility of WBE for a suite of enteric viral pathogens. We conduct a retrospective study to measure concentrations of enteric virus nucleic-acids in two samples each week of wastewater solids samples over 26 months from two wastewater treatment plants in the San Francisco Bay Area of California, US. In particular, we measured concentrations of human adenovirus group F, rotavirus, human norovirus GI and GII, and enterovirus nucleic-acids. These viruses were chosen for the study because they represent some of the most important etiologies of enteric viral infections globally^3,4^. In addition, we measured these targets in both the solids and liquid fractions of wastewater to assess their affinity to the solid matrix. Finally, we test whether wastewater concentrations of enteric virus nucleic-acids are associated with positivity rates from clinical samples from a local diagnostic laboratory.

## Methods

### (RT-)PCR assays

We used previously published assays for rotavirus^14^, human norovirus (HuNoV) GI^15^ and GII^16^, and enteroviruses (EV)^17^. We designed novel PCR primers and an internal hydrolysis probe for adenovirus group F (HAdV). HAdV genome sequences were downloaded from National Center for Biotechnology Information (NCBI) in April 2022 and aligned to identify conserved regions. Primers and probes were developed *in silico* using Primer3Plus (https://primer3plus.com/). Parameters used in assay development are provided in Table S1. All primers and probe sequences (Table S2) were screened for specificity *in silico* and *in vitro* against virus panels and genomic and synthetic target nucleic-acids (appendix p 2, Table S3). We also used previously published assays for the SARS-CoV-2^18^, and pepper mild mottle virus M gene (PMMoV)^2^.

### Study design and sample collection

This study is an observational, retrospective longitudinal surveillance study. Two WWTPs that serve 75% (1,500,000 people) of Santa Clara County, (SJ) and 25% (250,000 people) of San Francisco County (OSP), California were included in the study (Figure S1). Further WWTP descriptions are elsewhere^19^.

Wastewater solids samples were collected daily for a prospective WBE effort beginning November 2020 and stored, and a subset of those samples (two samples per week, 459 samples total) are used in this study. The samples were chosen to span a 26 month period (2/1/21 - 4/14/23) spanning different phases of the COVID-19 pandemic.

To compare concentrations of enteric virus nucleic-acids in liquid wastewater and wastewater solids, eight samples of settled solids and 24-hour composited influent were collected from OSP from the primary clarifier and the inlet to the plant, respectively, on July 11, 13-16, 17, 18, and 21, 2023 (n=8, hereafter labeled as samples 1-8) using clean sterile containers.

### Procedures

For the retrospective study, 50 mL of wastewater solids were collected using sterile technique in clean bottles from the primary clarifier. At SJ, twenty-four hour composite samples were collected by combining grab samples from the sludge line every six hours. At OSP, a grab sample was collected from the sludge line. Samples were stored at 4°C, transported to the lab, and processed within six hours. Solids were dewatered^20^ and frozen at - 80°C for 4 - 60 weeks. Frozen samples were thawed overnight at 4°C and then nucleic-acids were obtained from the dewatered solids following previously published protocols^18,21^. Nucleic-acids were obtained from 10 replicate sample aliquots. Each replicate nucleic-acid extract from each sample was subsequently stored between 8 and 273 days for SJ and between 1 and 8 days for OSP at −80°C and subjected to a single freeze thaw cycle. Upon thawing, targets were measured immediately using digital droplet (RT-)PCR with multiplexed assays for SARS-CoV-2, rotavirus, HAdV, HuNoV GI, HuNoV GII, EV, and PMMoV. PMMoV is highly abundant in human stool and wastewater ^22^ and is used as an internal recovery and fecal strength control. Each nucleic-acid extract was run in a single well so that 10 replicate wells were run for each sample for each assay.

In order to investigate potential losses from storage and freeze thaw of the samples and their nucleic-acid extracts, we compared measurements of the SARS-CoV-2 and PMMoV measured in the retrospective, longitudinal study obtained using the stored samples to measurements from the same samples that were not stored and analyzed prospectively for a regional monitoring effort (appendix p 3).

For the wastewater liquid/solid comparison, influent and solids samples were transported to the laboratory on ice and stored at 4°C for 7 days during which time we expect minimal decay of nucleic-acid target based on experiments with other similar targets^23^. The solids were dewatered and processed exactly as described above for the retrospective samples. The influent samples were processed to obtain nucleic-acid as described previously^10^; 10 subsamples were processed per sample (appendix p 2). Nucleic-acid templates were used immediately (no storage) as template in multiplexed (RT-)PCR reactions for rotavirus, HAdV, EV, HuNoV GI, HuNoV GII, and PMMoV. Each subsample nucleic-acid extract was run in a single well so that 10 replicate wells were run for each sample for each assay.

Further details of the dd(RT)-PCR, including the use of controls and thresholding, are described in the appendix (pp 2-3). We confirmed that multiplexing up to eight assays in a single reaction did not interfere with target quantification (appendix p 3). Concentrations of nucleic-acid targets were converted to concentrations in units of copies (cp)/g dry weight or per milliliter of liquid using dimensional analysis. The errors are reported as standard deviations. Three positive droplets across 10 merged wells corresponds to a concentration between ∼500-1000 cp/g; the range in values is a result of the range in the equivalent mass of dry solids added to the wells, or 1 cp/ml for liquid.

### Enteric virus infection positivity rate data

Data are not gathered on incidence and prevalence of rotavirus, adenovirus group F, or norovirus infections in the US. As such, we use rotavirus, adenovirus group F, or norovirus clinical test positivity rates to infer information about the spread of disease. We used positivity rates from Stanford Health Care Clinical Microbiology Laboratory (“clinical laboratory”) for comparison to wastewater data. The clinical laboratory tests stool specimens of symptomatic patients for rotavirus, adenovirus group F, and HuNoV using the BIOFIRE GI panel (bioMérieux, Inc., Salt Lake City, UT). HuNoV testing included both GI and GII, but is not disaggregated. The clinical laboratory is among the largest in Santa Clara County, where SJ WWTP is located, and receives specimens from counties in the San Francisco Bay Area. We assumed that the positivity rates recorded at the clinical laboratory are reasonable estimates for the positivity rates for residents in the sewersheds. Positivity rates are aggregated weekly. Enteroviruses are a diverse group of viruses and there is no clinical testing data available that represents disease occurrence caused by enteroviruses in aggregate.

### Statistical analysis

Data tended to be not normal, so non-parametric tests were used. We compared concentrations at SJ and OSP for each viral target using Mann-Whitney U tests (5 tests total). We tested the null hypothesis that infection positivity rates are not associated with the nucleic-acids concentrations in wastewater solids at each WWTP (8 tests total) using Kendall’s test of association; we used weekly aggregated positivity rate from the clinical laboratory, and average of the two weekly concentrations of viral nucleic-acids for the analysis. We tested the null hypothesis that nucleic-acid concentrations of each virus were not associated between the two WWTPs (5 tests total) using Kendall’s test; since samples from the exact same day at SJ and OSP were not processed, we used the weekly average concentrations. We tested the null hypothesis that viral nucleic-acid concentrations were not associated within each WWTP (20 tests total) using Kendall’s test. We used a p value of 0.0013 (0.05/38) for alpha=0.05 to account for multiple hypotheses testing. All analysis was carried out using RStudio (1.4.1106) and R (4.0.5).

### Ethics

This study was reviewed by theStanford University Institutional Review Board (IRB) and the IRB determined that this research does not involve human subjects and is exempt from oversight.

## Results

Results are reported as suggested in the Environmental Microbiology Minimal Information guidelines^24^ (appendix p 4, Figure S2, Table S4). Extraction and (RT-)PCR negative and positive controls performed as expected (negative and positive, respectively). In the 459 samples comprising the retrospective study, consistency of PMMoV measurements in samples within WWTP indicated no gross nucleic-acid extraction failures (appendix pp 3-4). Comparison of SARS-CoV-2 and PMMoV measurements made on unstored samples to those used in this retrospective study that were stored, and for which the nucleic-acid underwent one freeze-thaw gave equivocal results; PMMoV showed no degradation. However, median (IQR) ratios of measurements in stored and fresh samples was 0.2 (0.1-0.3) and 0.4 (0.2-0.6) for SJ and OSP, respectively, suggesting some degradation (appendix p 4). Although not ideal, storage of samples is essential for retrospective work.

Results of *in silico* analysis indicated no cross reactivity of the enteric virus (RT-)PCR probe-based assays with sequences deposited in NCBI; *in vitro* testing against non-target and target viral nucleic-acids (Table S3) also indicated no cross reactivity. We multiplexed eight different assays in each PCR reaction. We found that the concentration of each target was not affected by the presence of orthogonal targets as background (Figure S3).

Viral nucleic-acid concentrations in eight matched wastewater liquid and solid samples were orders of magnitude higher in the solids compared to the liquid wastewater, on a per mass basis (Figures 1 and S4). The distribution coefficients, K_d_, defined as the ratio of nucleic-acids concentrations in solids to liquids, in decreasing rank order of medians (IQR) are: EV K_d_ 26000 ml/g (12300-28700), HuNoV GI K_d_ 24400 ml/g (9800-28900), HuNoV GII K_d_ 16100 ml/g (10000-22400); HAdV K_d_ 6700 ml/g (I4600-13200); PMMoV K_d_ 3700 ml/g (3400-4400); and rotavirus K_d_ 650 (400-980). Given the observed enrichment of viral targets in wastewater solids, it is justified to use the solid matrix for prospective wastewater monitoring for infectious disease targets.

**Figure 1.**
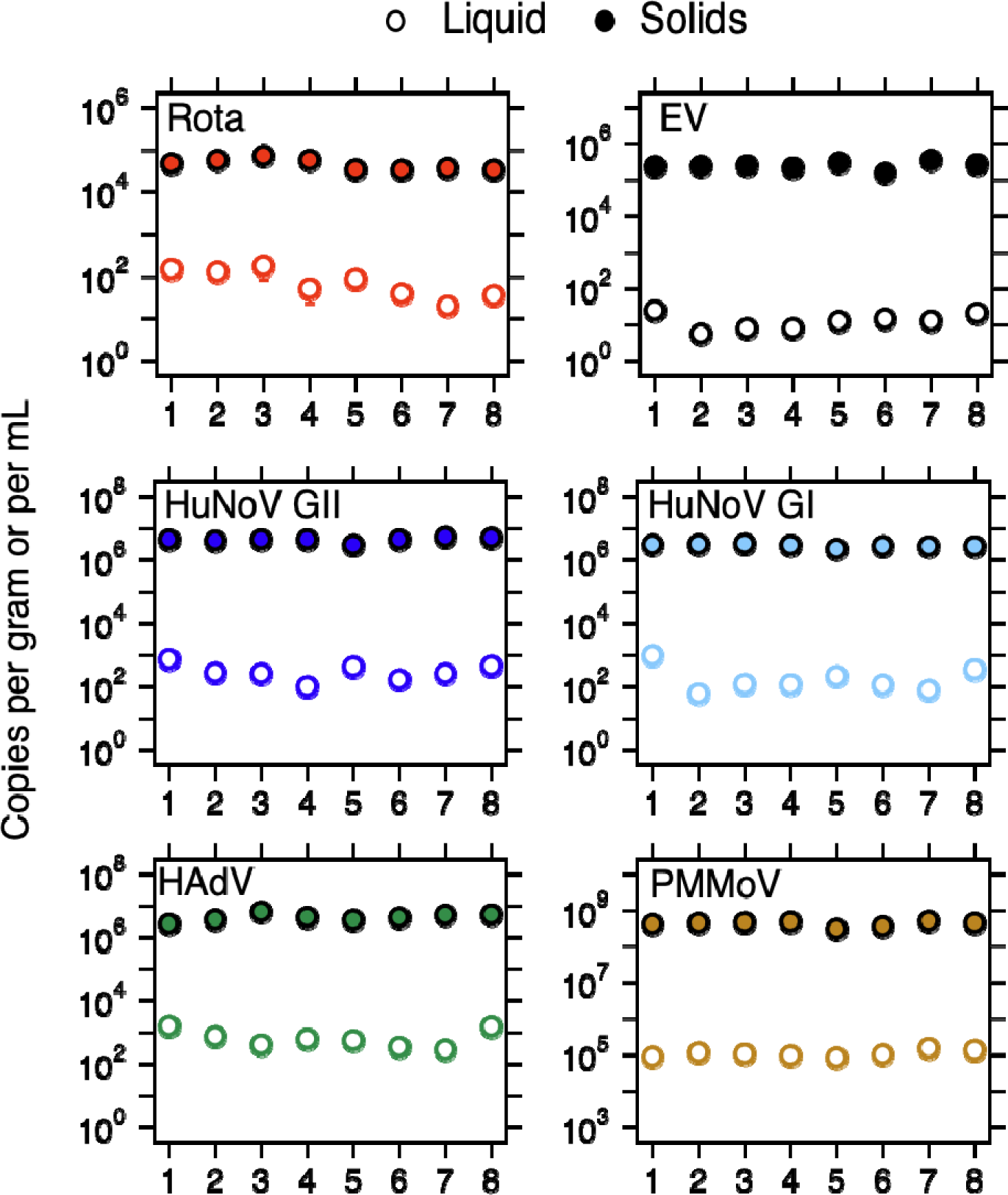
Concentration of viral targets in wastewater solids and liquid. Error bars are standard deviations and in most cases cannot be seen because they are smaller than the symbols. The x-axis shows the sample numbers (1-8) as indicated in the main text. The units on the y-axis are copies per gram of solids or milliliters of liquid. Rota is rotavirus, HAdV is human adenovirus group F, HuNoV GI and GII are human norovirus genogroup I and II, respectively, EV is enterovirus, and PMMoV is pepper mild mottle virus.

The retrospective, longitudinal study detected EV, HAdV, rotavirus, and HuNoV GI and GII nucleic-acids in wastewater solids (Figures 2 and S5) throughout the study with most showing some cyclic, season patterns with higher concentrations in winter/spring periods. Median concentrations of each enteric viral target were significantly higher at SJ relative to OSP (Mann-Whitney U Test, all p<0.001) for all viral targets except for HuNoV GII, but the effect size was less than 1 order of magnitude (Table 1). Within each WWTP, median concentrations of the different viruses ranked highest to lowest were HAdV (highest median concentration of 5.8×10^5^ and 1.9×10^6^ cp/g for OSP and SJ, respectively) followed by HuNoV GII (5.7×10^5^ and 7.0×10^5^ cp/g for OSP and SJ, respectively), EV (4.1×10^4^ and 5.4×10^4^ cp/g for OSP and SJ, respectively), HuNoV GI (2.8×10^4^ and 3.4×10^4^ cp/g for OSP and SJ, respectively), and rotavirus with the lowest (1×10^3^ and 3.3 x 10^3^ cp/g for OSP and SJ, respectively). Other summary statistics (interquartile ranges, maximums and number of non-detects) generally follow the same pattern (Table 1).

**Figure 2.**
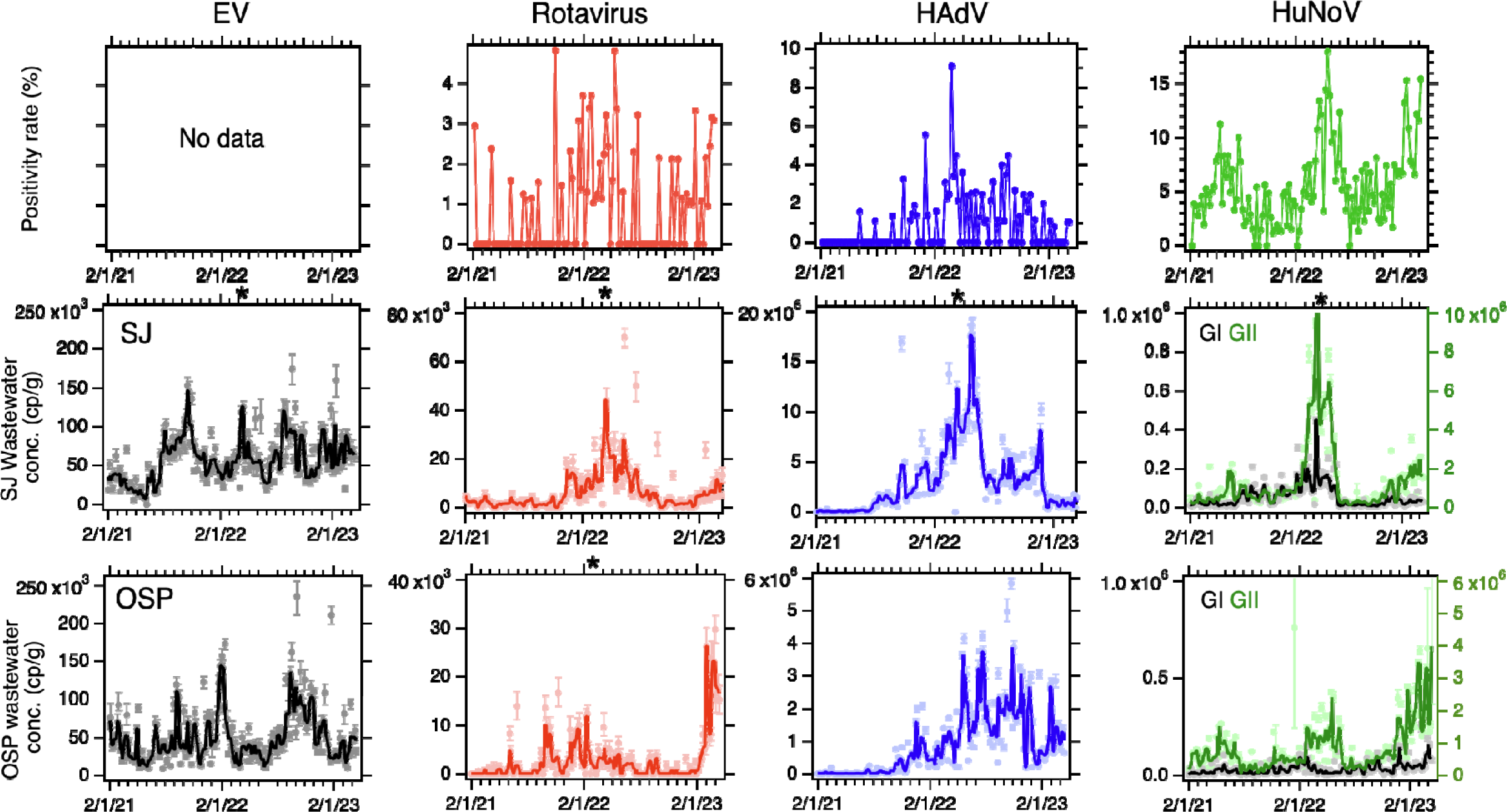
Positivity rates from the clinical laboratory for rotavirus, adenovirus group F, and norovirus infections (top panels), and concentrations of EV, rotavirus, HAdV, and HuNoV GI and GII nucleic-acids in wastewater solids at SJ (middle panels) and OSP (bottom panels). Error bars on the wastewater measurements show standard deviations. The solid lines in the wastewater plots represent smoothing using the median of 3-adjacent samples. The norovirus plots show GII in green (right axes) and GI in black (left axis). A black asterisk at the top of a plot indicates a value at that date that is higher than the y-axis scale. An asterisk on the top axis indicates a single data point out of range of the axis: EV at SJ: 1.2 x 10^6^ cp/g on 4/14/22, Rota at SJ: 2.7 x 10^5^ cp/g on 4/14/22, HAdV at SJ: 85 x 10^6^ cp/g on 4/14/22, HuNoV GI at SJ: 1.4 x 10^6^ cp/g on 4/14/22, HuNoV GII at SJ: 1 x 10^8^ cp/g on 4/14/22, and Rota at OSP: 78 x 10^3^ cp/g on 3/6/23.

**Table 1.** Summary statistics for infectious disease targets in OSP and SJ wastewater solids. WWTP is wastewater treatment plant. Units of concentration are copies per gram dry weight. 25th is 25th percentile, 75th is 75th percentile, max is the maximum value, # ND is number of non-detect, # is the total number of samples. Units for the concentrations are copies per gram dry weight.

| WWTP |  | Rota | HAdV | EV | HuNoV GI | HuNoV GII |
| --- | --- | --- | --- | --- | --- | --- |
| OSP | median | 1.0x10<sup>3</sup> | 5.8x10<sup>5</sup> | 4.1x10<sup>4</sup> | 2.8x10<sup>4</sup> | 5.7x10<sup>5</sup> |
|  | 25th | 0 | 4.9x10<sup>4</sup> | 2.6x10<sup>4</sup> | 1.2x10<sup>4</sup> | 3.5x10<sup>5</sup> |
| | 75th | $3.2 \times 10^3$ | $1.5 \times 10^6$ | $6.9 \times 10^4$ | $4.7 \times 10^4$ | $1.1 \times 10^6$ |
| | max | $7.8 \times 10^4$ | $5.8 \times 10^6$ | $2.4 \times 10^5$ | $2.6 \times 10^5$ | $6.4 \times 10^6$ |
|  | # ND | 89 | 9 | 0 | 0 | 0 |
|  | # | 230 | 230 | 230 | 230 | 230 |
| SJ | median | $3.3 \times 10^3$ | $1.9 \times 10^6$ | $5.4 \times 10^4$ | $3.4 \times 10^4$ | $7.0 \times 10^5$ |
| | 25th | $1.5 \times 10^3$ | $5.3 \times 10^5$ | $3.7 \times 10^4$ | $1.6 \times 10^4$ | $3.5 \times 10^5$ |
| | 75th | $8.5 \times 10^3$ | $4.3 \times 10^6$ | $7.5 \times 10^4$ | $7.6 \times 10^4$ | $1.5 \times 10^6$ |
| | max | $2.7 \times 10^5$ | $8.5 \times 10^7$ | $1.2 \times 10^6$ | $1.4 \times 10^6$ | $1.1 \times 10^8$ |
|  | # ND | 24 | 0 | 1 | 0 | 0 |
|  | # | 229 | 229 | 229 | 229 | 229 |

Between WWTPs, concentration of rotavirus, HAdV, and HuNoV GII nucleic-acids were positively associated (rotavirus tau = 0.27, p<10^−4^; HAdV tau = 0.49, p<10^−13^; HuNoV GII tau = 0.36, p<10^−7^) whereas HuNoV GI and EV concentrations were not correlated between WWTPs. Within SJ WWTP, the viral nucleic-acid concentrations were positively associated with each other (tau between 0.25 and 0.6 depending on viral target, all p < 10^−4^) except that EV was not associated with rotavirus, HuNoV GII, or HuNoV GI nucleic-acid concentrations. Within OSP, rotavirus and HuNoV GI (tau = 0.23, p = 0.00036), HuNoV GII and GI (tau = 0.30, p<10^−5^), and HAdV and EV (tau = 0.21, p =0.00079) nucleic-acid concentrations were positively associated.

Time series of enteric virus nucleic-acids in relation to SARS-CoV-2 RNA measured herein, and other respiratory virus nucleic-acids reported at SJ previously^7^ are provided in Figure 3. At SJ, enteric virus nucleic-acid concentrations increased after the BA.1 surge starting on 2/1/22 and peaked contemporaneously near the BA.2 surge peak, and declined sharply after the BA.2 surge. This is in contrast to the pattern observed for a suite of non-SARS-CoV-2 respiratory viruses (including parainfluenza, metapneumovirus, influenza, and coronaviruses) that peaked just prior to the BA.1 surge, and decreased sharply after, and then rose again during the BA.2 surge. At OSP, a local maxima in the enteric virus nucleic-acid concentrations occurs near the BA.2 surge, as observed at SJ, but thereafter, patterns for the different viruses diverge. HAdV and HuNoV GI remained relatively high for the duration of the time series after the BA.2 surge, while rotavirus and HuNoV GII concentrations were highest at the end of the time series (spring 2023).

**Figure 3.**
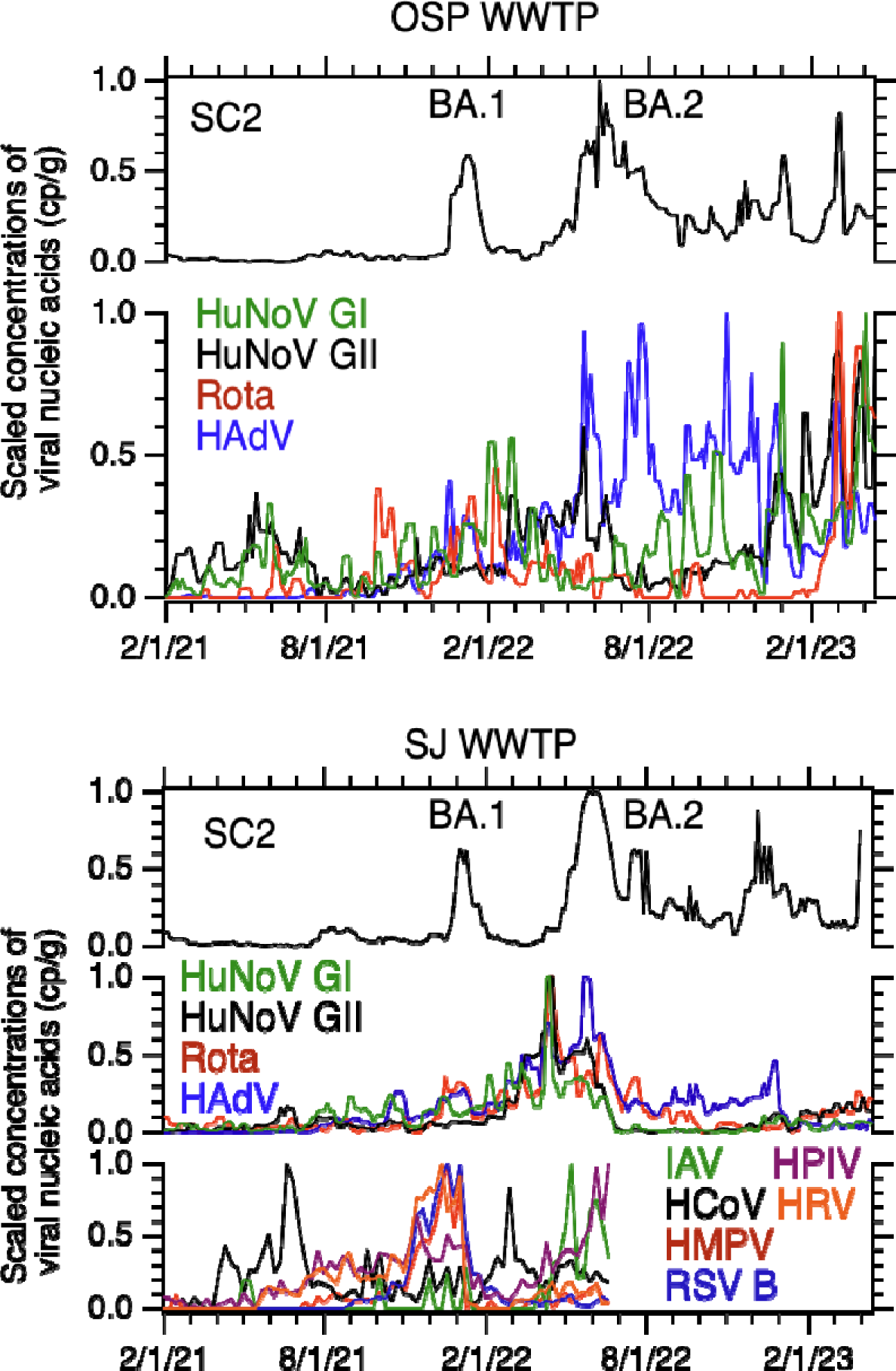
Standardized virus concentrations. Standardized, 3-sample smoothed (using median) concentrations of the HuNoV GI and GII, rotavirus (Rota), and human adenovirus (HAdV) (middle or bottom left axis) and SARS-CoV-2 (SC2, top axis) measured in this study in wastewater solids from OSP (top plot) and SJ (bottom plot). The BA.1 and BA.2 surges in SC2 concentrations is labeled in each plot atop of adjacent, respectively, to the surges. Data from respiratory viruses previous published by Boehm et al.^7^ at SJ through mid summer 2022 are provided on the bottom of the SJ plot; IAV is influenza A, HCoV is seasonal coronaviruses, HMPV is human metapneumovirus, HPIV is human parainfluenza virus, HRV is human rhinovirus and RSV B is respiratory syncytial virus B. The dominant SARS-CoV-2 variant circulating is provided at the top of the figure near each SARS-CoV-2 surge in concentration. The standardized concentration (C_sd_) is calculated using the following formula C_sd_(t) = C(t) - C_min_/(C_max_ - C_min_) where C(t) is the smoothed concentration of the viral target, and C_max_ and C_min_ are the max and min of the smoothed concentrations observed over the duration of the time series.

The clinical laboratory processed a median (IQR) of 80 (66-89) samples per week . Median (IQR) positivity rates were lowest for rotavirus (0% (0-1.4%), max = 4.8%), intermediate for adenovirus group F (0 (0 −1.5%), max = 9.1%) and highest for HuNoV (4.8% (2.8-7.5%), max = 18.0%). HAdV in wastewater solids at both WWTPs was positively correlated to adenovirus positivity rates (tau = 0.30 for OSP and 0.38 for SJ, both p<10^−4^). HuNoV GII in wastewater solids at both WWTPs was positively correlated to norovirus positivity rates (tau = 0.36 for OSP and 0.26 for SJ, both p<10^−4^). HuNoV GI in wastewater solids was not associated with norovirus positivity rates (tau = 0.16, p = 0.014 for OSP, tau = 0.0067, p = 0.92 for SJ). Rotavirus in wastewater solids at SJ was positively associated with rotavirus positivity rates (tau = 0.24, p=0.00049); however, rotavirus at OSP was not associated with positivity rate (tau = 0.19, p = 0.0081).

## Discussion

We detected rotavirus, human adenovirus group F (HAdV), enterovirus (EV), and human norovirus (HuNoV) genogroups I and II in wastewater solids at two San Francisco Bay Area WWTPs twice per week for 26 weeks. We observed coherence of the viral target concentrations between the two WWTPs, suggesting some regional coherence in community infections and shedding into wastewater. Many of the targets showed temporal coherent patterns within WWTPs with maxima in concentrations in the winter/spring seasons. Viral enteric infections are usually seasonal with higher rates in the winter months^25^, and seasonal wastewater patterns are consistent with that despite various behavior changes potentially associated with the COVID-19 pandemic over the course of the study. Temporal variation of the enteric viral targets are distinct from those of SARS-CoV-2 and other respiratory viruses in wastewater, suggesting divergent disease dynamics. However, low enteric virus nucleic-acid concentrations in the winter/spring of 2021, relative to the following years may be a result of non-pharmaceutical interventions in place during the early period of COVID-19 pandemic, particularly prior to 4/14/21 when vaccines became widely available, which may have limited the spread of enteric viral infections.

Concentrations of rotavirus, HAdV, and HuNoV GII in wastewater solids were associated with clinical positivity rates for infections associated with the viruses. This is despite the biases and limitations associated with the clinical data; specifically that the tests were administered on specimens from severely ill individuals, and the individuals tested may not reside or work in the sewersheds of the two WWTPs. While there is no ground-truth data available to determine disease occurrence, results suggest that enteric viral nucleic-acid concentrations in wastewater solids can yield insight into enteric disease activity in contributing communities. The lack of correlation of HuNoV GI and norovirus positivity rates is consistent with data suggesting HuNoV GII is the main genogroup causing symptomatic illness in this region^26^. The EV assay used in this study is a pan-*Enterovirus* genus assay and therefore detects a number of different viruses including coxsackieviruses, enteroviruses, and echoviruses. The diversity of viruses detected with the assay, and the diversity of presentations associated with them, including meningitis, diarrhea, rash, and respiratory symptoms means that there is no clinical metric that can be used for comparison to the collected wastewater data. However, detection of these viruses in wastewater suggests occurrence of enterovirus infections in the communities.

Concentrations of enteric virus nucleic-acid targets are enriched by orders of magnitude in wastewater solids relative to liquid on mass equivalent basis. These findings are consistent with previous reports that viruses in wastewater tend to adsorb to wastewater solids^12^. There was variation in the degree of adsorption among viruses suggesting virus structure might influence adsorption characteristics. Ye et al.^27^ suggest enveloped viruses partition more favorably to wastewater solids than non-enveloped viruses, but our findings suggest non-enveloped viruses are enriched in wastewater solids to a similar extent^13^, at least when they are measured using molecular methods. There are limited data on concentrations of enteric virus nucleic-acids in wastewater solids^12^, and to our knowledge, no longitudinal measurement of a suite of enteric viruses measured contemporaneously. However, some of our results are consistent with observations made in liquid wastewater. For example, HAdV is reported to be among the highest concentration enteric viruses in wastewater^28^, and herein, it was found at the highest concentrations in wastewater solids.

Rotavirus was detected at the lowest concentration of enteric viruses tested, consistent with the fact that live, oral rotavirus vaccines are regularly administered to infants in the study area and the resultant low clinical positivity rates. Based on available rotavirus vaccine sequences, the rotavirus assay may detect vaccine genomic RNA. As infants in our study area typically wear diapers, it is unclear to what extent the vaccine strains from infants may appear in wastewater. However, as vaccine strains are live, transmission to others who do use the wastewater system may be possible, with their shedding possibly influencing concentrations of rotavirus RNA in the waste stream.

Worldwide there are limited to no direct data on the incidence and prevalence of enteric viral infections, and the data that do exist are biased to those who are severely ill or to individuals who seek and obtain diagnostic testing. Wastewater represents a composite biological sample of the entire contributing community and concentrations of enteric virus nucleic-acids can provide insights into disease occurrence including occurrence of asymptomatic and mild enteric virus infections. Such information can inform clinical decision making, hospital staffing, or informational alerts to the public about the circulation of disease. Additional work at WWTPs is warranted to substantiate these findings. Further work is needed to better characterize viral shedding characteristics to inform model-based estimates of disease incidence and prevalence from wastewater measurements^29^.

## Data Availability

Wastewater data are available publicly at the Stanford Digital Repository (https://doi.org/10.25740/vx726fw9373). Positivity rate data for clinical specimens are available upon request.

## Acknowledgements

We acknowledge the numerous people at San Jose and Oceanside wastewater treatment plants who contributed to wastewater sample collection and Allegra Koch for her support with literature review. This research was performed on the ancestral and unceded lands of the Muwekma Ohlone people. We pay our respects to them and their Elders, past and present, and are grateful for the opportunity to live and work here.

## Data sharing statement

Wastewater data are available publicly at the Stanford Digital Repository (https://doi.org/10.25740/hr647tm4528). Positivity rate data for clinical specimens are available upon request.

## Author contributions

Alexandria Boehm: conceptualisation, data curation, formal analysis, funding acquisition, investigation, methodology, project administration, resources, software, supervision, validation, visualisation, writing – original draft, writing – review & editing. Bridgette Hughes: conceptualisation, data curation, formal analysis, investigation, methodology, writing – original draft, writing – review & editing. Dorothea Duong: conceptualisation, data curation, formal analysis, investigation, methodology, writing – review & editing. Niaz Banaei: data curation, methodology, validation, writing – review & editing. Bradley White: conceptualisation, data curation, investigation, methodology, project administration, resources, software, supervision, validation, writing – review & editing. Marlene Wolfe: conceptualisation, project administration, writing – review & editing.

