## Appendix for "A retrospective longitudinal study of adenovirus group F, norovirus GI and GII, rotavirus, and enterovirus nucleic-acids in wastewater solids at two wastewater treatment plants: Solid-liquid partitioning and relation to clinical testing data"

Contents (ToC heading absent)

**Supplementary Methods** **2**

**Supplementary Results** **3**

Table S1. Parameters used in assay development software. Parameters used in the development of new primers and probes using Primer3Plus (https://primer3plus.com/, accessed 7/16/23). 6

Table S2. Primers and probes used in this study for different targets. 7

Table S3. Viruses considered in this study and the region of the genome that each virus assay targets. Viruses used to test specificity are indicated as “non target testing” and viruses used as positive controls are indicated as “target testing”. 8

Table S4. Additional details related to the EMMI guidelines. 9

Figure S1. The location of the two sewersheds where samples were collected in this study. 10

Figure S2. EMMI15 checklist. 11

Figure S3. Multiplex assay performance. 12

Figure S4. Distribution of Kd values measured in eight samples of paired liquid and solid samples from OSP for each viral target considered herein. 13

Figure S5. PMMoV-normalized wastewater data and positivity rates. 14

### Supplementary Methods

**(RT-)PCR assays.** All primers and probe sequences (including custom and previously published assays, Table S2) were then screened for specificity *in silico* and *in vitro*. Assays were tested in vitro for specificity and sensitivity using virus panels (NATtrol™ Respiratory Verification Panel NATRVP2.1-BIO, NATtrol™ EV Panel NATRVPEVP-C, Zeptometrix, Buffalo, NY), and genomic and synthetic target nucleic acids purchased from American Type Culture Collection (ATCC, Manassas, VA) (Table S3). The respiratory virus panel includes chemically inactivated intact influenza viruses, parainfluenza viruses, adenovirus, rhinovirus A, metapneumovirus, rhinovirus, RSV, several coronaviruses, and SARS-CoV-2; the EV panel includes chemically inactivated intact coxsackieviruses, echovirus, and parechovirus. Nucleic acids were extracted from intact viruses or cells using Chemagic Viral DNA/RNA 300 Kit H96 for Chemagic 360 (PerkinElmer, Waltham, MA).

Nucleic acids were used undiluted as template in digital droplet (RT-)PCR singleton assays for sensitivity and specificity testing in single wells. The concentration of targets used in the in vitro specificity testing was between $10^3$ and $10^4$ copies per well. Negative (RT-)PCR controls were included on each plate.

**Solids pre-analytical processing.** As mentioned in the main text, these methods have been described in detail previously in published papers[1,2] and open access protocols[3,4]. Briefly, a pre-measured mass of dewatered solids was suspended in a buffer at a concentration of 75 mg/ml. The mixture was homogenized and then centrifuged; nucleic-acids were extracted using a commercial kit Chemagic Viral DNA/RNA 300 Kit H96 for Chemagic 360 (PerkinElmer, Waltham, MA) and then the nucleic-acids were processed using an inhibitor removal kit (Zymo, Irvine, CA). This protocol has been shown to alleviate potential inhibition while maintaining good assay sensitivity[2,5]. 300 µl of the suspension entered into the nucleic-acid extraction process and 50 µl of nucleic-acids are retrieved after the inhibitor removal kit. An aliquot of the dewater solids was used to determine the dry weight of the solids using oven drying.

**Influent pre-analytical processing.** For each influent sample, 10 replicate aliquots were processed using an affinity-based capture method with magnetic hydrogel Nanotrap Particles with Enhancement Reagent 1 (Ceres Nanosciences, Manassas, VA) on 10 mL of sample to concentrate viral particles using a KingFisher Flex system following vendor instructions. Nucleic-acids were then extracted from the each concentrated aliquot using the MagMAX Viral/Pathogen Nucleic Acid Isolation Kit (Applied Biosystems, Waltham, MA) on the KingFisher Flex platform to obtain purified nucleic acids which were then process through a Zymo OneStep-96 PCR Inhibitor Removal kit (Zymo Research, Irvine, CA). Each 10 ml sample resulted in 50 µl total nucleic acid extract.

**dd(RT-)PCR methods.** The ddRT-PCR methods applied to measure PMMoV in a singleplex reaction are provided in detail elsewhere[1]. The human virus assays were run in multiplex using a probe-mixing approach and unique fluorescent molecules (HEX, FAM, Cy5, Cy5.5, ROX, ATTO950) in two sets of reactions. One reaction included primers and probes for rotavirus (fluorescent molecule(s) on probe: FAM), SARS-CoV-2 (FAM/HEX), HuNoV GII (ATTO590), and HAdV (ROX/ATTO590). The second reaction included primers and probes for EV (Cy5.5) and HuNoV GI (ATTO590). Note that the two human virus reactions contained several additional primers and probe sets that yielded results not reported herein. In particular, assays targeting the genomes of West Nile virus, human immunodeficiency virus, hepatitis A virus, *Candida auris*, and enterovirus D68 and several influenza subtype markers[6] were included. Each reaction was run on its own 96-well plate.

Each 96-well PCR plate of wastewater samples included PCR positive controls for each target assayed on the plate in 1 well, PCR negative no template controls in two wells, and extraction negative controls (consisting of water and lysis buffer) in two wells. PCR positive controls consisted of viral gRNA or gene blocks (Table S3).

ddRT-PCR was performed on 20 µl samples from a 22 µl reaction volume, prepared using 5.5 µl template, mixed with 5.5 µl of One-Step RT-ddPCR Advanced Kit for Probes (Bio-Rad 1863021), 2.2 µl of 200 U/µl Reverse Transcriptase, 1.1 µl of 300 mM dithiothreitol (DTT) and primers and probes mixtures at a final concentration of 900 nM and 250 nM respectively. Primer and probes for assays were purchased from Integrated DNA Technologies (IDT, San Diego, CA) (Table 1). Human virus targets were measured in reactions with undiluted template whereas PMMoV was run on template diluted 1:100 in molecular grade water.

Droplets were generated using the AutoDG Automated Droplet Generator (Bio-Rad, Hercules, CA). PCR was performed using Mastercycler Pro (Eppendforf, Enfield, CT) with with the following cycling conditions: reverse transcription at 50°C for 60 minutes, enzyme activation at 95°C for 5 minutes, 40 cycles of denaturation at 95°C for 30 seconds and annealing and extension at 59°C (for human viruses) or 56°C (for PMMoV) for 30 seconds, enzyme deactivation at 98°C for 10 minutes then an indefinite hold at 4°C. The ramp rate for temperature changes were set to 2°C/second and the final hold at 4°C was performed for a minimum of 30 minutes to allow the droplets to stabilize. Droplets were analyzed using the QX200 or the QX600 Droplet Reader (Bio-Rad). A well had to have over 10,000 droplets for inclusion in the analysis. All liquid transfers were performed using the Agilent Bravo (Agilent Technologies, Santa Clara, CA).

Thresholding was done using QuantaSoft™ Analysis Pro Software (Bio-Rad, version 1.0.596) and QX Manager Software (Bio-Rad, version 2.0). Replicate wells were merged for analysis of each sample. In order for a sample to be recorded as positive, it had to have at least 3 positive droplets.

We sought to confirm that the multiplexing up to eight assays did not interfere with target quantification. We tested whether the quantification of a single target in the presence of and absence of similar concentrations of the seven other targets, including those for which results are not reported in this study, was substantially different. To do so, we first quantified three decimal dilutions of a target nucleic acid in the absence of any other targets using the (RT)-PCR chemistry that included primers and probes for all eight targets. Then, we quantified the same decimal dilutions of the single target in the presence of 10-100 copies per reaction of the seven other targets. Each dilution was run in a single well and no template, negative controls were included on each PCR plate. Assays were run and thresholded as described above. Results were expressed as copies per reaction and the standard deviation, as output by the instrument, were included.

**Comparison of PMMoV and SARS-CoV-2 measurements on stored versus unstored samples.** All the samples used in this study were processed for measurements of SARS-CoV-2 N gene and PMMoV M gene in wastewater solids on unstored samples as a part of a prospective wastewater monitoring program for infectious disease surveillance. The methods for analysis for the N gene through 3/13/23 were identical to those previously described in a Data Descriptor[2] with the exact same approaches described in this paper except the assay was multiplexed with two other assays using a two color droplet reader (QX200, Bio-rad). Between 3/13/23 and 4/14/23, the N gene was multiplexed with assays for five adjacent single nucleotide polymorphisms (SNPs) in SARS-CoV-2 XBB*, respiratory syncytial virus (RSV), influenza A, influenza B, and HuNoV GII in conjunction with a 6-color droplet reader (QX600, Bio-rad)[6–8].

### Supplementary Results

**PMMoV measurements in the samples used for the retrospective analysis.** Since PMMoV RNA is present in high concentrations in the samples naturally, lack of its detection, or abnormally low measurements might indicate gross extraction failures. The median (interquartile range) $\log_{10}$-transformed PMMoV was 9.1 (9.0-9.2) and 8.7 (8.6-

8.8) $\log_{10}$ copies/g at SJ and OSP, respectively. The lowest measurements at the two sites were 8.5 (SJ) and 8.1 (OSP) $\log_{10}$ copies/g and given these lowest values are within an order of magnitude of the medians, we concluded that there was no gross extraction failure. We opted to not use an exogenous viral control in these experiments, like bovine coronavirus, owing to the complexities associated with interpreting recovery of an exogenous spiked control in environmental samples[9], and its potential to interfere with other uses of the samples, for example viral metagenomics.

**Losses attributable to storage and freeze thaw.** We compared measurements of the SARS-CoV-2 N gene and PMMoV M gene made on the samples that were fresh to those used in this study that were stored, and for which the RNA underwent one freeze-thaw. Median (IQR) ratio of PMMoV measurements made in this study to those made using fresh samples was 0.9 (0.6-1.2) at SJ and 1.0 (0.7-1.4) at OSP suggesting limited degradation of the PMMoV target in the stored and freeze-thawed samples. Median ratio of SARS-CoV-2 N gene measurements made in this study to those made using fresh samples was 0.2 (0.1-0.3) at SJ and 0.4 (0.2-0.6) at OSP suggesting storage and freeze thaw may have reduced measurement concentrations, but by less than an order of magnitude.

**Additional details related to the EMMI guidelines.** Thirty-six samples from the retrospective study were selected at random for this analysis; this represents 8% of the samples processed in the study. As described in the methods section, each sample was run as template in three different PCR reactions; 1 for PMMoV, 1 for rotavirus, SARS-CoV-2, HuNoV GII, and HAdV, and 1 for EV and HuNoV GI. The average (standard deviation) number of partitions (droplets) for each of the three reactions (across the 10 replicates) was 164164 (38851) for the reaction for PMMoV, 174321 (20126) for the reaction for rotavirus, SARS-CoV-2, HuNoV GII, and HAdV, and 185497 (24410) for the reaction for EV and HuNoV GI. The volume of the partitions, as reported by the machine vendor is 0.00085 µL. The mean and standard deviation of copies per partition for each target is shown in Table S4. Example fluorescent plots from the QX200 (two color reader) can be viewed in Topol et al. on protocols.io[10] and an example fluorescent plot from the QX600 (6 color reader) is included in the Stanford Digital Repository with the deposited data (https://doi.org/10.25740/hr647tm4528).

**Table S1. Parameters used in assay development software. Parameters used in the development of new primers and probes using Primer3Plus (https://primer3plus.com/, accessed 7/16/23).**

- Product size ranges: 60-275
- Primer size: min 15 bp, opt 20 bp, max 36 bp
- Primer melting temperature: min 50°C, optimal 60°C, max 65°C
- GC% content: min 40%, optimal 50%, high 60%
- concentration of divalent cations = 3.8 mM
- concentration of dNTPs needs to be 0.8 mM
- Internal Oligo: size min 15 bp, optimal 20 bp, max 30 bp
- Internal Oligo: Melting temp min 62°C, optimal 63°C, max 70°C
- Internal Oligo: GC% min 30%, optimal 50%, max 80%

| Target | Primer/Probe | Sequence |
|---|---|---|
| HAdV (183bp) | Forward | CCTCCTGTGTTACGCCAGA |
| | Reverse | CAGGCTGAAGTASGTATCGG |
| | Probe | CTCGATGATGCCGCAATGGT |
| HuNoV GII (88bp) | Forward | ATGTTCAGRTGGATGAGRTTCTCWGA |
| | Reverse | TCGACGCCATCTTCATTCACA |
| | Probe | AGCACGTGGGAGGGCGATCG |
| HuNoV GI (76bp) | Forward | GCCATGTTCCGITGG ATG |
| | Reverse | TCCTTAGACGCCATCATCAT |
| | Probe | TG GGACAGGAGATCGCAATCTC |
| Rotavirus (113bp) | Forward | CAGTGGTTGATGCTCAAGATGGA |
| | Reverse | TCATTGTAATCATATTGAATACCCA |
| | Probe | ACAACTGCAGCTTCAAAAGAAGWGT |
| EV (143bp) | Forward | CCCTGAATGCGGCTAAT |
| | Reverse | TGTCACCATAAGCAGCCA |
| | Probe | ACGGACACCCAAAGTAGTCGGTTC |
| SARS-CoV-2 (143bp) | Forward | CATTACGTTTGGTGGACCCT |
| | Reverse | CCTTGCCATGTTGAGTGAGA |
| | Probe | CGCGATCAAAACAACGTCGG |
| PMMoV | Forward | GAGTGGTTTGACCTTAACGTTTGA |

| | | |
|---|---|---|
| (68bp) | Reverse | TTGTCGGTTGCAATGCAAGT |
| | Probe | CCTACCGAAGCAAATG |

**Table S2. Primers and probes used in this study for different targets. The size of the amplicon generated by the primers is shown under the target name (units of basepairs, bp). Primers and probes were purchased from Integrated DNA Technologies (Coralville, IA, USA). All probes contained fluorescent molecules and quenchers (5′ HEX, FAM, Cy5, Cy5.5, ROX, and/or ATTO950/ZEN/3′ IBFQ); FAM, 6-fluorescein amidite; HEX, hexachloro-fluorescein; Cy5, Cyanine-5; Cy5.5, Cyanine5.5; ROX, carboxyrhodamine; ZEN, a proprietary internal quencher from Integrated DNA Technologies (Coralville, IA, USA); and IBFQ, Iowa Black FQ. HAdV is human adenovirus group F, HuNoV is human norovirus, and EV is enterovirus.**

| Virus | Genomic target | Non-target testing (negatives) | Target testing (positives) | Reference |
|---|---|---|---|---|
| Adenovirus F (HAdV) | Hexon gene | NATRVP2.1-BIO | ATCC VR-930DQ | Not applicable |
| Norovirus GII (HuNoV GII) | ORF1-2 junction | NATRVP2.1-BIO | ATCC VR-3235SD | Loisy et al. [11] |
| Norovirus G1 (HuNoV GI) | ORF1-2 junction | NATRVP2.1-BIO NATEVP-C | ATCC VR-3234SD | Jothikuman et al. [12] |
| Rotavirus | Non-structural Region Protein 3 (NSP3) | NATRVP2.1-BIO NATEVP-C | ATCC VR-2018DQ | Jothikumar et al. [13] |
| Enterovirus (EV) | 5' UTR | NATRVP2.1-BIO | NATEVP-C | Gregory et al.[14] |

**Table S3. Viruses considered in this study and the region of the genome that each virus assay targets. Viruses used to test specificity are indicated as "non target testing" and viruses used as positive controls are indicated as "target testing". The reference for each assay is provided aside from HAdV for which an assay was developed herein. All non-target controls are panels sold by Zeptomatrix (panels begin with NAT prefix, "Zepto", Buffalo, NY). ATCC is American Type Culture Collection. The NATRVP2.1-BIO panel includes chemically inactivated intact influenza viruses, parainfluenza viruses, adenovirus, rhinovirus, metapneumovirus, and coronaviruses. The NATEVP-C panel includes chemically inactivated intact coxsackieviruses, echovirus, and parechovirus. The full list of species in the panels is available from the vendor.**

**Table S4. Additional details related to the EMMI guidelines. For each target measured in this study, the mean and standard deviation (sd) of the total number of copies of target per partition. Num is the number of samples out of a random 36 included in this analysis that had detectable target in them and thus contributed to the calculated mean and standard deviation. A value of 0 indicates that of the random 36 samples selected, none of them had the target present in them. Abbreviations for the targets are provided in the main text except "Rota" is rotavirus and SC2 is the N gene of SARS-CoV-2.**

| Target | EV | HuNoV GI | Rota | SC2 | HuNoV GII | HAdV | PMMoV |
|---|---|---|---|---|---|---|---|
| mean | $1.02x10^{-3}$ | $8.93x10^{-4}$ | $1.20x10^{-4}$ | $1.37x10^{-3}$ | $1.12x10^{-2}$ | $5.03x10^{-2}$ | 0.15 |
| sd | $3.65x10^{-4}$ | $7.94x10^{-4}$ | $1.14x10^{-4}$ | $1.22x10^{-3}$ | $8.57x10^{-3}$ | $3.58x10^{-2}$ | 0.069 |
| num | 36 | 36 | 23 | 36 | 36 | 36 | 36 |

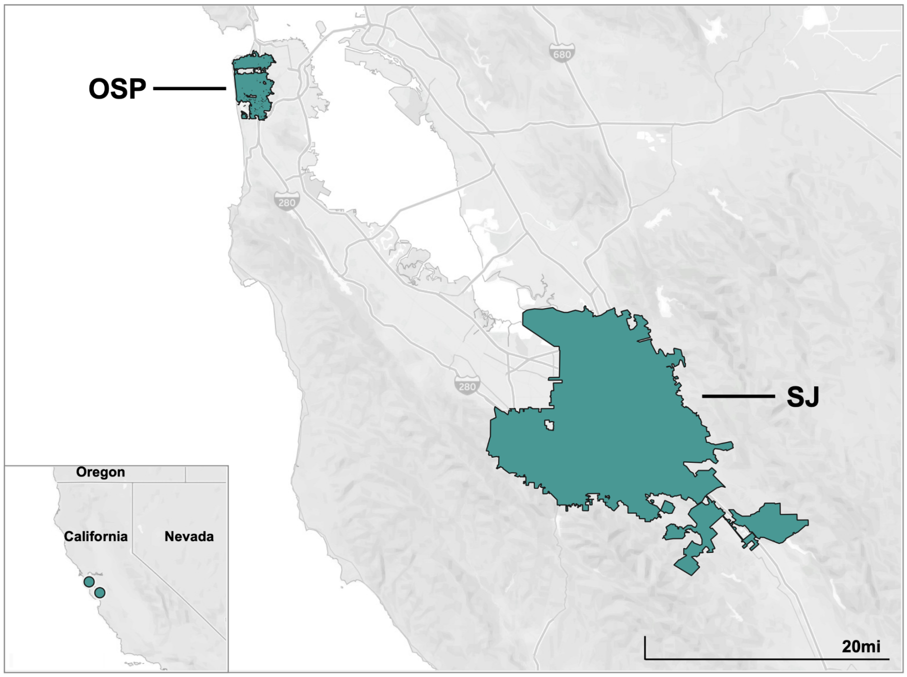


**Figure S1. The location of the two sewersheds where samples were collected in this study.**

**Figure S2. EMMI[15] checklist. The checklist is also available at the Stanford Digital Repository with the deposited data (https://doi.org/10.25740/hr647tm4528).**

| Study Description | Environmental Sampling | Sample Treatment | Sample Reduction | Nucleic-acid Extraction | Reverse Transcription | PCR Amplification | Analysis |
|---|---|---|---|---|---|---|---|
| Study name: enteric virus<br>Date: 8/17/23<br>Completed by: Boehm | Notes: Described in detail in the methods section | Notes: No sample treatment done | Notes: Described in detail in the methods section | Notes: Described in detail in the methods section | Notes: Described in detail in the methods section | Notes: Described in detail in the methods section | Notes: Described in detail in the methods section |

| Control Checklist | Environmental Sampling | Sample Treatment | Sample Reduction | Nucleic-acid Extraction | Reverse Transcription | PCR Amplification | |
|---|---|---|---|---|---|---|---|
| Step performed | ☑ | ☐ | ☑ | ☑ | ☑ | ☑ | |
| Step has control info | ☐ | ☐ | ☐ | ☐ | ☐ | ☐ | Negative controls |
| # of control replicates | 0 | na | 0 | 2 | 2 | 2 | |
| Control result reported | ☐ | ☐ | ☐ | ☑ | ☑ | ☑ | |
| Method for handling failed controls described | ☐ | ☐ | ☐ | ☑ | ☑ | ☑ | |
| Step has control info | ☐ | ☐ | ☑ | ☑ | ☑ | ☑ | Positive controls |
| Control identity described | ☐ | ☐ | ☑ | ☑ | ☑ | ☑ | |
| Control quantification method described | ☐ | ☐ | ☑ | ☑ | ☑ | ☑ | |
| # control replicates | 0 | na | 10 | 1 | 1 | 1 | |
| Control result reported | ☐ | ☐ | ☑ | ☑ | ☑ | ☑ | |
| Method for handling failed controls described | ☐ | ☐ | ☑ | ☑ | ☑ | ☑ | |

Process checklist

| Environmental Sampling | |
|---|---|
| Sample procedure | ☑ |
| Number of samples | ☑ |
| Sample amount, mean, range | ☑ |
| Sampling locations, dates, times | ☑ |
| Sample storage conditions | ☑ |

| Sample Treatment | |
|---|---|
| Treatment procedure | ☐ |
| Reagents | ☐ |

| Sample Reduction | |
|---|---|
| Reduction procedure | ☑ |
| Reagents | ☑ |
| Concentration factor | ☑ |

| Nucleic-acid Extraction | |
|---|---|
| Extraction procedure | ☑ |
| Volume or mass extracted, volume or mass obtained | ☑ |
| Extract storage conditions | ☑ |

| Reverse Transcription | |
|---|---|
| One- or two-step | ☑ |
| cDNA storage conditions (if 2 step) | ☐ |
| Reaction temperatures and times | ☑ |
| Reaction reagents and concentrations | ☑ |
| Priming method | ☑ |
| Reaction volume, added template amount | ☑ |
| RT efficiency assessment procedure (if 2-step) | ☐ |
| RT control description (if two-step) | ☐ |
| RT efficiency reported (if 2-step) | ☐ |

| qPCR or dPCR | |
|---|---|
| Target gene name, amplicon length | ☑ |
| Thermocycling temp and times | ☑ |
| Master mix composition: vendors, concentrations | ☑ |
| Additives: vendors, composition | ☑ |
| Template amount added, pre-treatment (if any) | ☑ |
| Primers: sequences, concentrations, vendors, references | ☑ |
| Amplicon confirmation method (probe, melt curve details, etc) | ☑ |
| Probe sequence, concentration, vendor, reference | ☑ |
| Instrumentation | ☑ |
| Inhibition assessment procedure | ☑ |
| Inhibition control description (if used) | ☐ |
| Number of samples tested and found inhibited | ☑ |

| Analysis- dPCR | |
|---|---|
| Threshold settings | ☑ |
| Technical replicates, number, well merging | ☑ |
| Partitions measured, number, mean, variance | ☑ |
| Partition volume | ☑ |
| Target copies per partition, mean, variance | ☑ |
| Program used for dPCR analysis | ☑ |
| Explanation of control results, example plots | ☑ |

| Analysis- qPCR | |
|---|---|
| Technical replicates, number, calculations | ☐ |
| Calibration standards, description, source | ☐ |
| Method of quantifying standards | ☐ |
| Calibration curve slope | ☐ |
| Calibration curve R2 | ☐ |
| Lowest standard measured or 95% LOD | ☐ |
| Cq value determination methods | ☐ |

Note to users: This checklist is provided as guidance for best practices for reporting, but is not meant to be prescriptive. Not all items in the check list will apply to all studies. Please see Borchardt et al. The Environmental

Version 2.0

This version maintained by Borchardt, Boehm, Salt, Noble, Wiggington, Spencer

Date: 4 August 2023

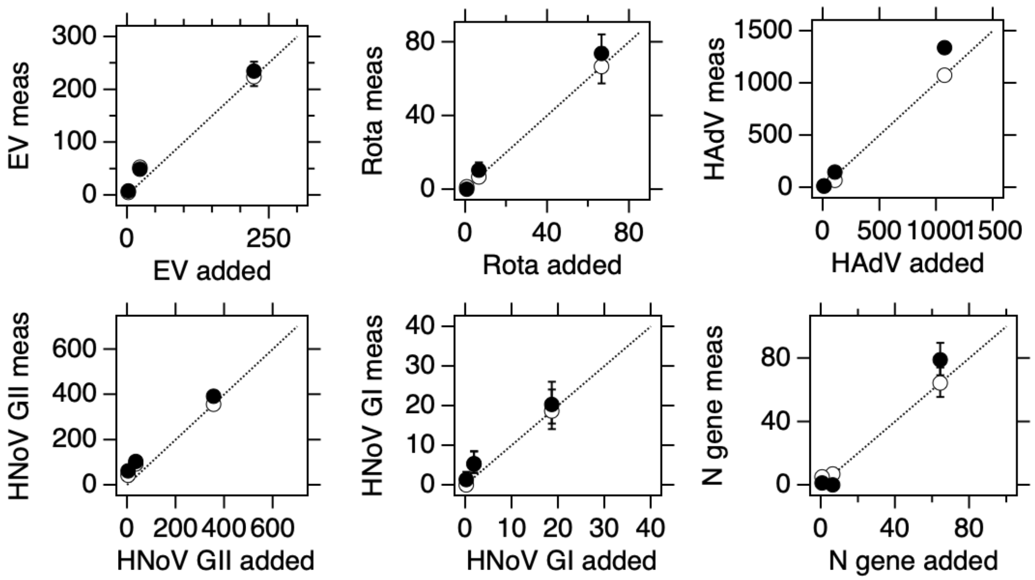


**Figure S3. Multiplex assay performance. The concentrations of the different targets in units of copies per reaction input and measured in each experiment is provided. The white symbols are for reactions without the 7 background nucleic-acid targets and black symbols include high concentrations of the 7 other nucleic-acid targets. Error bars are standard deviations, if error bars cannot be seen, then they are smaller than the symbol. The line represents the 1:1 line. "Meas" is measured, "Rota" is rotavirus. Abbreviations for the different targets are provided in the main text.**

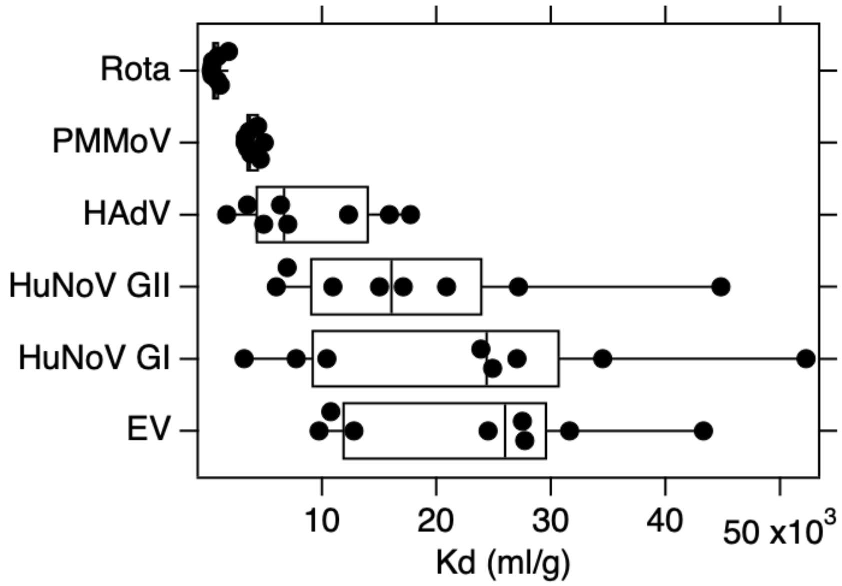


**Figure S4. Distribution of Kd values measured in eight samples of paired liquid and solid samples from OSP for each viral target considered herein. The points represent each of the eight $K_d$ values. The midline of each box is the median and the edges of the box represent the 25th and 75th percentiles. The whiskers extend to the 9th and 91st percentiles. Rota is rotavirus, and the remaining abbreviations are used in the text of the paper. The order of the viruses from top to bottom of the y-axis is smallest to largest median $K_d$.**

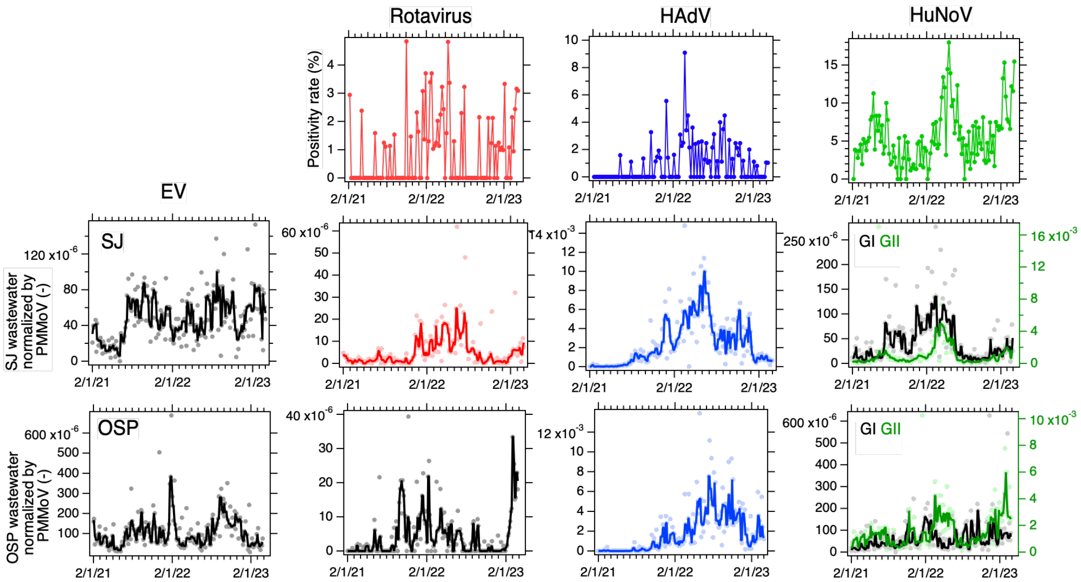


**Figure S5. PMMoV-normalized wastewater data. Positivity rates from the clinical laboratory for rotavirus, adenovirus group F, and norovirus infections (top panels), and concentrations of EV, rotavirus, HAdV, and HuNoV G1 and GII nucleic-acids in wastewater solids at OSP (middle panels) and SJ (bottom panels) normalized by concentrations of PMMoV measured in the sample. The solid lines in the wastewater plots represent smoothing using the median of 3-adjacent samples. The norovirus plots show GII in green (right axes) and GI in black (left axis).**
